# Multi-trait and Gene-Based Analyses Identify Genetic Variants Associated with Spontaneous Coronary Artery Dissection

**DOI:** 10.64898/2026.03.12.26348247

**Authors:** Takiy-Eddine Berrandou, Adrien Georges, Ingrid Tarr, Eleni Giannoulatou, Robert M Graham, Doug Speed, Nabila Bouatia-Naji

**Author notes:** CORRESPONDING AUTHOR: Nabila Bouatia-Naji, PhD, Paris Cardiovascular Research Centre, 56, rue Leblanc, 75015, PARIS, FRANCE.

## Abstract

**Background and aims:** Spontaneous coronary artery dissection (SCAD) is a non-atherosclerotic cause of acute myocardial infarction (MI) that predominantly affects young women. As an under-recognized cause of MI, large genome-wide association studies (GWAS) remain challenging. We aimed to leverage SCAD shared genetic basis with related vascular diseases to uncover genetically determined biological mechanisms.

**Methods:** Summary statistics for SCAD GWAS (1,917 cases, 9,293 controls) was harmonised with seven related vascular traits: fibromuscular dysplasia, intracranial aneurysm, cervical artery dissection, migraine, coronary artery disease, abdominal aortic aneurysm, and thoracic aortic aneurysm/dissection. We applied Multi-Trait Analysis of GWAS (MTAG). We integrated coronary-artery regulatory annotations, cis-eQTL mapping, and colocalization to prioritize candidate genes. Gene-based testing (LDAK-GBAT) was applied to SCAD dataset.

**Results:** MTAG identified 40 independent SCAD loci, including 24 that were novel. Candidate variants were enriched in open chromatin from coronary smooth muscle cells and fibroblasts and in vascular regulatory regions. LDAK-GBAT identified 46 significant genes, including 12 outside MTAG loci. Integrated functional annotation prioritized 56 genes linked to arterial integrity, vasoactive tone, haemostasis, and coagulation. Extracellular matrix organization was confirmed as a key pathway, with additional enrichment in bone mineralization and TGF-β related terms.

**Conclusions:** Integrating multi-trait GWAS, gene-based testing, epigenetic and transcriptomic data substantially expanded the SCAD genetic landscape. Our findings implicate key arterial-wall pathways beyond extracellular matrix organization, and point at relevant biological mechanisms in non-atherosclerotic dissection. These findings nominate tractable targets for experimental follow-up and support future efforts toward SCAD risk stratification in women.

## INTRODUCTION

Spontaneous coronary artery dissection (SCAD) is recognized as a non-atherosclerotic cause of acute myocardial infarction that predominantly affects women, often in the absence of conventional cardiovascular risk factors. SCAD accounts for a substantial proportion of myocardial infarctions in women younger than 50 years and of pregnancy-associated infarctions ^1–5^. SCAD is defined as a non-traumatic, non-iatrogenic separation of the coronary artery wall caused by an intimal tear or bleeding from the vasa vasorum, leading to intramural hematoma formation, luminal compression and myocardial ischemia ^1,3,5,6^. Observational cohorts indicate that SCAD remains underdiagnosed, carries a substantial risk of recurrent events, and shows a strong association with extra-coronary arteriopathies, particularly fibromuscular dysplasia (FMD). Other non-atherosclerotic vascular phenotypes are also commonly reported in SCAD, such as cervical artery dissection, intracranial aneurysm, and migraine ^1,5–10^. Familial clustering and co-occurrence of SCAD with Mendelian connective-tissue and aortic disorders is rare, as most SCAD cases are sporadic, with an incompletely understood pathophysiology ^4,11–13^. A notable subset of SCAD events occurs during pregnancy or postpartum, suggesting that genetic susceptibility acts in a vascular context shaped by sex-specific hormonal and hemodynamic states ^1,5,6,14^.

Recent genetic efforts support SCAD as a polygenic disease. The largest SCAD genome-wide association study (GWAS) meta-analysis to date reported 16 significant loci, near genes involved mostly in extracellular-matrix biology^13^. Consistent with its clinical overlap with extra-coronary arteriopathies, SCAD shares several of its GWAS loci with fibromuscular dysplasia, cervical artery dissection, and intracranial aneurysm, in addition to a negative genetic correlation with atherosclerotic coronary artery disease^13^. These observations suggest that additional SCAD loci are likely to be shared with most of these vascular diseases and traits, yet remain undetected in single-trait GWAS.

The under-diagnosed nature of SCAD due to lack of systematic angiography exploration near the index cardiac event renders it challenging to improve genetic discovery by enlarging SCAD numbers, even in the context of international consortia ^1,5,9,13,15^. This contrasts with atherosclerotic coronary artery disease, where large biobanks enable orders-of-magnitude larger GWAS and hundreds of loci have already been identified using these resources ^16^. These constraints motivate the application of approaches that borrow information from genetically correlated vascular traits while preserving the specific inference to SCAD phenotype.

To expand locus discovery while retaining SCAD-centred inference, we jointly analysed SCAD with seven genetically related vascular traits using a multi-trait GWAS framework ^17^ and complementary leave-one trait-out analyses. We also applied gene-based association testing to the original SCAD dataset to capture loci driven by an aggregation of sub-threshold variant effects to help improve discovery power ^18^. Integrated arterial-wall regulatory annotations, tissue specific transcriptomics and eQTL data from GTEx, and colocalization were applied to prioritize genes and pathways to enhance biological translation from novel genetic findings^19,20^.

## MATERIAL and METHODS

### GWAS datasets and quality control

We assembled genome-wide association summary statistics from eight traits related to vascular and neurovascular biology to conduct multi-trait analyses centred on spontaneous coronary artery dissection (SCAD). Detailed information about each dataset, including sample sizes and sources, is provided in Supplementary Table 1.

SCAD summary statistics were obtained from our most recent meta-analysis ^13^, including 1,917 cases and 9,293 controls of European ancestry. FMD summary statistics were obtained from our GWAS meta-analysis of fibromuscular dysplasia ^21^. Additional summary statistics were drawn from GWAS meta-analyses of intracranial aneurysm (IA) ^22^, cervical artery dissection (CeAD) ^23^, migraine, coronary artery disease (CAD) ^24^, abdominal aortic aneurysm (AAA) ^25^ and thoracic aortic aneurysms and dissections (TAAD) ^26^. For IA and CeAD, summary statistics were shared directly by the original investigators, whereas datasets for migraine, CAD, AAA and TAAD were obtained from public GWAS repositories as listed in Supplementary Table 1.

All summary statistics were subjected to the same quality control (QC) pipeline. We retained only variants meeting the following criteria: minor allele frequency (MAF) > 0.01; imputation quality score (INFO or Rsq) > 0.9; Hardy-Weinberg equilibrium (HWE) p-value > 1 × 10⁻⁶; and biallelic status. For binary traits, the effective sample size (Neff) was calculated using the formula: N_eff_=4/(1/N_Cases_+1/N_Controls_). This harmonized dataset of high-quality variants served as the basis for all subsequent multi-trait and gene-based analyses.

### Imputation of missing summary statistics

To harmonize genome-wide association summary statistics across all traits and increase variant coverage, we imputed missing Z-scores using the GAUSS R package (v1.0.0) ^27^. GAUSS performs ancestry-informed imputation using a reference panel of 32,953 genomes (the 33KG panel), including over 20,000 European samples. For each trait, we supplied formatted summary statistics including chromosome, position, alleles, and Z-scores. The imputation was performed using the “dist()” function under the assumption of European ancestry, using genomic windows defined by linkage disequilibrium (LD) blocks from the LAVA algorithm, which partitions the hg19 genome into 2,495 regions of approximately 1 Mb while minimising linkage disequilibrium between blocks (median width 1.02 Mb) ^28^.

Post-imputation quality control retained only biallelic variants with an imputation INFO score ≥ 0.8 and removed duplicate rsIDs. For each trait, GAUSS-imputed Z-scores were added only for SNP–trait combinations absent from the original GWAS summary statistics; all observed GWAS estimates were retained unchanged. This procedure modestly increased per-trait SNP counts for well-covered GWAS (e.g., SCAD from 6.70 to 6.93 million SNPs; Supplementary Table 2) and increased the number of SNPs shared across all eight traits from 3.58 million (pre-imputation) to approximately 4.85 million (post-imputation). As a quality check to avoid reliance on imputed SCAD statistics for discovery, we confirmed that the reported lead variants were present in the original SCAD summary statistics.

### Multi-trait GWAS Analysis

We applied Multi-Trait Analysis of GWAS (MTAG) ^17^ to jointly analyse SCAD with seven genetically correlated traits: FMD, IA, CeAD, migraine, atherosclerotic CAD, AAA and TAAD (Supplementary Figure 1). Given effect size estimates from multiple GWAS, MTAG computes a revised set of effect size estimates for the target trait (here, SCAD). The weightings assigned to each trait are designed to minimize the squared difference between the revised estimates and the true effect sizes, and depend on the GWAS sample sizes, the genetic correlations between pairs of traits, and the sample overlap between the corresponding GWAS (MTAG estimates the genetic correlations and sample overlaps using LDSC).

MTAG is implemented as a generalized least-squares estimator. For each SNP_j_, the vector of marginal effect estimates across traits is modelled as the sum of a latent vector of true genetic effects and sampling error, assuming a constant trait-by-trait covariance of per-SNP effects across the genome. The required genetic covariance matrix and SNP-specific sampling covariance matrices are estimated from summary statistics using LD score regression, which enables MTAG to correct for unknown and potentially substantial sample overlap. We ran MTAG using the default settings and LD score regression options recommended by the original authors.

### Panel of MTAG configurations and selection-adjusted inference

To reduce sensitivity of SCAD locus discovery to the inclusion of any single auxiliary trait, we implemented a pre-specified panel of eight MTAG configurations, each producing SCAD-specific association statistics. This panel comprised (i) the full model including SCAD and all seven auxiliary traits, and (ii) seven leave-one-out (LOO) configurations, each excluding one auxiliary trait (FMD, IA, CeAD, migraine, CAD, AAA, or TAAD) while retaining SCAD and the remaining six traits. For locus discovery, we summarized evidence across the eight correlated MTAG configurations using a single selection-adjusted p-value, P*_sel_*, which explicitly accounts for selecting the most extreme SCAD association signal across the panel ^29,30^.

For each SNP_j_, let *Z_r_* denote the SCAD Z-statistic from MTAG configuration r ∈ {1, …, 8}, and define Y = *max_r_* = 1.8|Z*_r_*|. Under the null hypothesis of no association, the vector (*Z*_1_, …, *Z*_8_) is approximately multivariate normal with mean 0 and correlation matrix *R* induced by the shared SCAD data and the overlap in auxiliary-trait sets across configurations. We estimated *R* empirically using LD-pruned HapMap3 variants, excluding SNPs with *max_r_*|*Z_r_*| ≥ 3 to limit contamination by true signals. We then computed 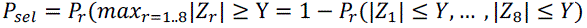, which corresponds to an adjustment for the minimum P-value across correlated tests ^29^. P*_sel_* was obtained by numerical evaluation of the multivariate normal probability over [−*Y*, *Y*]^8^ using Genz’s algorithm as implemented in the R package mvtnorm ^31–33^. For computational efficiency, P*_sel_* was obtained by interpolation from a precomputed grid of Y values, with a conservative Bonferroni upper bound used for extreme Y. Unless stated otherwise, all variant-level results and locus-discovery thresholds refer to P*_sel_*; we declared genome-wide significance at P*_sel_* < 5 × 10^-8^.

### Locus definition and lead-variant selection

SNPs were grouped into loci by physical distance (±500 kb around the most significant variant). Within each locus, the lead SNP was defined as the variant with the smallest selection-adjusted P-value P*_sel_*. For each lead SNP, we report P*_sel_* and the corresponding MTAG Z-statistic from the MTAG configuration that achieved the maximum |*Z*| (equivalently, the minimum configuration-specific p-value) used to construct P*_sel_*. To ensure consistency with the original SCAD data, we required the direction of effect of the lead SNP to be concordant between the selected MTAG run and the SCAD-only GWAS, but we did not impose an additional P-value threshold in the SCAD-only analysis given its limited power.

Unless stated otherwise, summary statistics and figures referring to “SCAD MTAG” results correspond to this union of 8 traits MTAG and MTAG-LOO analyses, using the selection-adjusted P*_sel_* for inference. In particular, on the Manhattan plots, for each SNP, P*_sel_* derived from the maximum |*Z_r_*| across the eight MTAG configurations. Regional association plots were generated using LocusZoom ^34^ to visualize lead SNPs and their local linkage disequilibrium context.

### Validation in an independent SCAD case control study

We evaluated lead variants in an independent SCAD case–control dataset from the **V**ictor Chang Cardiac Research Institute **A**rteriopathies & **S**CAD **C**ohort (VASC). After cohort-level quality control, the dataset comprised 1127 controls and 481 SCAD cases (359 array-genotyped and 122 whole-genome sequenced). Analyses were restricted to individuals of European ancestry defined by principal-component analysis (460 cases, 1127 controls). Array genotypes underwent standard variant-level QC (Hardy–Weinberg equilibrium P > 1×10⁻⁴, call rate > 99%, MAF > 0.005) and relatedness filtering using KING (kinship threshold 0.177). Array data were imputed to the HRC r1.1 European reference panel and variants were filtered to imputation r² ≥ 0.8. Whole-genome sequence data were restricted to PASS variants and filtered using matched thresholds, followed by cohort-level relatedness filtering. Association testing used Firth logistic regression implemented in PLINK v2, adjusting for sex, the first five principal components, and an array-versus-WGS indicator. To minimize overlap with the discovery meta-analysis, we report primary look-up results in a non-overlapping case subset excluding individuals previously included in the SCAD meta-GWAS (293 cases, 1127 controls), with sensitivity analyses in the full European-ancestry case set (460 cases, 1127 controls).

### Gene-based association analysis

We conducted gene-based association testing using SCAD meta-analysis GWAS summary statistics (harmonized as described above) and the LDAK-GBAT framework ^18^. LDAK-GBAT uses a linear mixed model to estimate the heritability of each gene (the proportion of phenotypic variation explained by the SNPs in the gene), then derives a gene-based p-value by testing whether the estimated heritability is significantly greater than zero. LDAK-GBAT requires GWAS summary statistics and a reference panel (the latter is used to estimate SNP-SNP correlations, should have similar ancestry to the GWAS).

For this analysis, we used an external European-ancestry LD reference panel of 10,000 unrelated UK Biobank participants ^35^ with imputed genotypes (hg19 build), comprising 6,901,602 SNPs (the overlap between those in the SCAD summary statistics and those in the UK Biobank with information score >=0.8)Gene definitions were based on the Ensembl v105 RefSeq annotation (NCBI37.3 genome build) ^36^ without additional upstream or downstream extensions. Among 19,427 annotated autosomal genes, 17,926 contained at least one SNP and were included in the analysis, corresponding to 2,750,177 unique variants.

We ran LDAK-GBAT using “Human Default Model”, as recommended by the authors ^18^, where SNP heritability is weighted by MAF according to the function [MAF_j_(1−MAF_j_)]^0.75^. Genome-wide significance was defined using a Bonferroni threshold of P < 2.79 × 10⁻⁶, based on the number of tested genes. To reduce redundancy among highly correlated gene signals, we applied clumping based on estimated genetic contributions: genes with squared correlation (r²) > 0.1 on the same chromosome were grouped, and only the most strongly associated gene in each group was retained.

### Genomic reference and gene models

All genomic coordinates are reported on the genome build of the original GWAS summary statistics (GRCh37/hg19 unless otherwise stated).

### Variant annotation and regulatory enrichment

At each SCAD locus, we defined candidate functional variants as the union of (i) variants belonging to the 95% credible set, estimated with the “ppfunc” function of the corrcoverage R package (v1.2.1), within ±500 kb of the lead SNP, and (ii) variants in high linkage disequilibrium (LD; r² > 0.8) with the lead SNPs in European-ancestry samples from the 1000 Genomes Project, identified using the “ldproxy” function in LDlinkR (v1.4.0) ^37^. Variant consequences were retrieved from Ensembl using the “biomaRt” package (v2.62.1) ^38^.

Enrichment of candidate variants in open chromatin and active regulatory regions was assessed with GREGOR (v1.4.0) ^39^, with parameters set to exclude any LD proxies (LDWINDOWSIZE = 1). Histone H3K27ac ChIP–seq peak files were downloaded from ENCODE (https://www.encodeproject.org/). Peak files corresponding to the same tissue (biosample term name) were merged using bedtools (v2.30.0). Coronary artery single-nucleus ATAC–seq (snATAC–seq) raw reads were obtained from the Sequence Read Archive (SRP321576) and processed following the original analysis pipeline to regenerate pseudo-bulk peak files representative of each cell cluster ^19^. The complete list of regulatory datasets used is provided in Supplementary Table 3.

### eQTL lookup and colocalization

To investigate the impact of SCAD variants on gene expression, we queried significant cis-eQTLs from GTEx ^40^ v10 release (www.gtexportal.org/home/datasets) in ten tissues relevant to vascular and cardiometabolic biology: tibial nerve, cultured fibroblasts, tibial artery, coronary artery, aorta, whole blood, liver, lung, heart (left ventricle) and heart (atrial appendage). From this, eQTLs overlapping with lead SNPs and candidate functional variants at SCAD MTAG loci were identified.

For colocalization analyses, we downloaded all SNP–gene cis-eQTL summary statistics from the GTEx v10 release. Colocalization between SCAD MTAG association signals and eQTLs was evaluated using the coloc R package (v5.2.3) with default prior settings ^20^. We considered there to be evidence for colocalization when the posterior probability for a shared causal variant (H4) exceeded 0.8. For each colocalized locus, the top candidate SNP was defined as the variant minimising the product of the eQTL and SCAD MTAG association P-values. LD between this top candidate and surrounding variants was obtained from the European 1000 Genomes reference panel using the LDproxy function in LDlinkR (v1.4.0) ^37^. Multi-trait colocalization was performed using HyprColoc R package (v1.0) ^41^.

### Gene prioritization and functional annotation

Genes were prioritized using five complementary lines of evidence: (i) proximity to SCAD-associated lead SNPs identified in the original SCAD meta-analysis GWAS or the MTAG analysis, defined as the nearest gene, (ii) significant gene-based association in LDAK-GBAT (gene-based association methods are described in a separate section) (iii) significant cis-eQTLs involving candidate variants, (iv) genetic colocalization between SCAD MTAG and eQTL signals, and (v) presence of at least one missense variant among the SCAD candidate variants at the locus. For each locus, we prioritized genes that carried missense variants when applicable and were supported by colocalization or displayed the strongest gene-based association. When no gene fulfilled these criteria, we selected as the prioritized gene the one with the most lines of supporting evidence across the six categories.

Pathway enrichment analyses were conducted on prioritized genes using the clusterProfiler R package (v4.14.6) ^42^. Prioritized genes were further annotated with human disease associations from the ClinVar database (accessed October 2024) and mouse phenotypes from the Mouse Genome Informatics database. Enrichment of disease terms among SCAD-associated genes was assessed using the “enricher” function in clusterProfiler. In downstream tables, we indicate for each prioritized gene whether it is linked to enriched diseases, any ClinVar disease annotations, enriched pathways or mouse cardiovascular phenotypes.

### Data visualization tools and methods

Unless otherwise stated, plots were generated in R (v4.4.2) using the following packages: ggplot2 (v4.0.0), rtracklayer (v1.66.0), ggrepel (v0.9.6), dplyr (v1.1.4), tidyr (v1.3.1), readr (v2.1.5) and colorspace (v2.1-1).

## RESULTS

### Multi-trait analysis expands the SCAD association landscape

To increase power for locus discovery while retaining SCAD-specific inference, we first confirmed that SCAD is genome-wide genetically correlated with each of the auxiliary traits (Supplementary Figure 1). In addition to previously reported correlations with FMD, IA, CeAD, migraine, and CAD, we observed positive genetic correlation with AAA and TAAD (Supplementary Figure 1). We therefore implemented a pre-specified panel of eight MTAG configurations (full 8-trait model plus seven leave-one-out configurations) and used the selection-adjusted p-value (*P_sel_*) as the primary inference for variant-level discovery.

Across this MTAG panel, we identified 40 independent loci reaching genome-wide significance (P*_sel_* < 5 × 10^)(^), of which 24 have not been previously reported for SCAD (Figure 1, Supplementary Table 4). Thirty loci were genome-wide significant in the full 8-trait MTAG configuration, whereas ten loci achieved genome-wide significance only in at least one leave-one-out configuration, indicating that at some loci SCAD association signals are more detectable when a specific auxiliary trait is excluded. For example, a previously reported locus on chr1p21.3 near *F3* did not reach genome-wide significance in the full configuration but became significant in a leave-one-out configuration. In the full 8-trait MTAG configuration, the increase in mean χ² corresponded to a GWAS-equivalent effective sample size increase for SCAD from 6,356 to 9,354 individuals (cases and controls), representing an approximate 47% gain in statistical power relative to the original SCAD GWAS.

**Figure 1.**
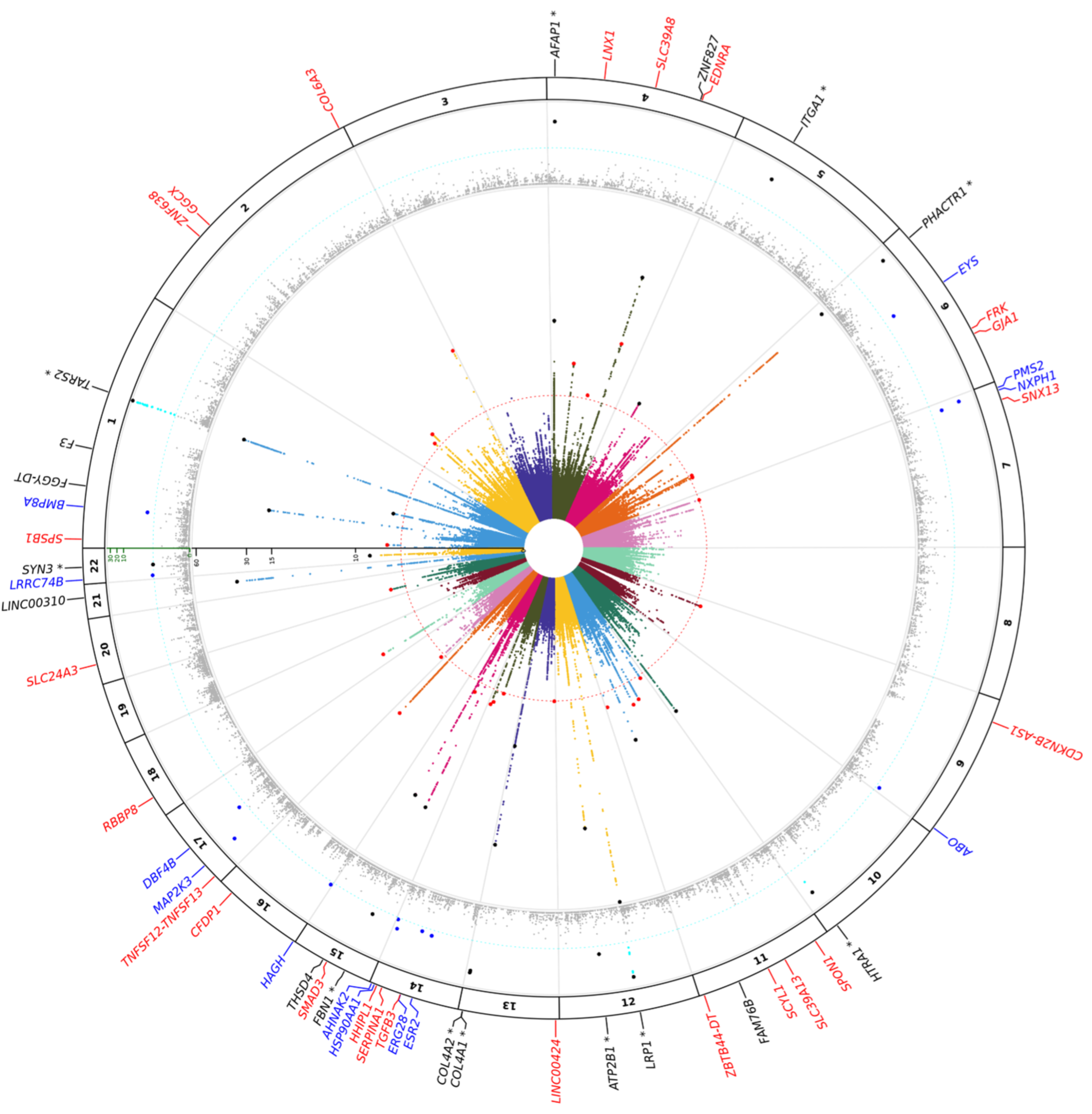
Integrated genomic landscape of spontaneous coronary artery dissection (SCAD). The circular plot summarizes variant-level multi-trait association and gene-based association results across autosomes (chromosomes 1–22; GRCh37/hg19). The innermost ring shows a radial Manhattan plot of SCAD associations from multi-trait GWAS analysis. The radial axis is *-log_10_ (P_sel_)*, where *P_sel_* is the selection-adjusted P-value accounting for the maximum evidence of association observed across eight correlated MTAG configurations (one primary eight-trait model including fibromuscular dysplasia, intracranial aneurysm, cervical artery dissection, migraine, coronary artery disease, abdominal aortic aneurysm, and thoracic aortic aneurysm/dissection, plus seven leave-one-out iterations). The red dashed circle marks genome-wide significance (*P_sel_* = 5 × 10^-8^). Novel MTAG loci are highlighted in red, and previously reported SCAD loci in black. The middle ring displays gene-based association results from LDAK-GBAT (linear mixed model) for 17,925 protein-coding genes, plotted at gene midpoint coordinates. The cyan dashed circle marks the Bonferroni threshold (*P < 2.79 × 10^-6^*); 46 genes exceed this threshold and are shown in cyan. Among Bonferroni-significant genes, independent lead genes (LD-clumped at r^2^ > 0.1) are emphasized: blue (n = 12) indicates novel GBAT signals located >1 Mb from any MTAG-significant SNP locus, whereas black (n = 10) indicates significant genes that fall within an MTAG-significant SNP locus. The outer labels annotate prioritized loci. For MTAG loci, the closest gene to each independent lead SNP is shown (red for novel loci, black for known loci). Blue labels denote novel GBAT lead genes. Loci marked with an asterisk (*) indicate convergence, where both MTAG (variant-level) and GBAT (gene-based) analyses independently reach their respective genome-wide significance thresholds.

All 24 novel loci showed the same direction of effect for all MTAG configurations and in the SCAD-only GWAS. Seven out of 24 showed only suggestive evidence of association (P ≤ 1 × 10⁻^5^; min= 8.3 × 10⁻⁷, max=9.5 × 10⁻²) in the recent SCAD GWAS meta-analysis (Supplementary Figure 2, Supplementary Table 5). Inspection of the cross-trait association patterns and leave-one-out MTAG results indicated that most of the 24 novel loci were supported across multiple MTAG configurations rather than being driven by a single auxiliary trait, although some showed stronger configuration dependence, particularly the signal near *CDKN2B-AS1*. Overall, MTAG reinforced, rather than reversed, the direction of SCAD effects observed in the SCAD-only GWAS (Supplementary Figures 2–3, Supplementary Table 5).

### Look up of MTAG-novel loci in an-independent SCAD case control cohort

To assess independent support for the MTAG association findings, while recognizing that the available sample size was insufficient for formal single-variant replication, we evaluated the association of the lead variants at the 24 novel loci in an independent SCAD case-control study using Firth logistic regression adjusted for sex, the first five ancestry principal components, and genotyping platform (array versus WGS). In the primary non-overlapping sample excluding cases included in the discovery SCAD meta-analysis (293 cases, 1,127 controls), 17 of 24 lead variants showed directionally consistent effects (binomial sign test *P* = 0.032), providing directional support for the MTAG findings at the polygenic level despite limited power for locus-by-locus replication. Three loci also showed nominal association with concordant direction (LNX1, SMAD3, and CFDP1; *P* < 0.05). Results were similar in a sensitivity analysis including all SCAD cases from this study (460 cases, 1,127 controls), with directionally consistent effects for 19 of 24 MTAG-novel variants (*P* = 0.0033), while acknowledging partial sample overlap with the discovery SCAD GWAS dataset ^13^ (Supplementary Table 6)

### Gene-based testing identifies additional SCAD loci beyond single-variant MTAG

We aggregated variant effects at the gene level with LDAK-GBAT ^18^, which models the joint heritable contribution of all SNPs annotated to a gene (see Methods). Using SCAD meta-analysis summary statistics and LD estimates from a 10,000-participant European UK Biobank reference panel, we tested 17,926 autosomal coding genes. Forty-six genes exceeded the Bonferroni threshold for genome-wide significance (*P* < 2.79 × 10^−6^; Figure 1, Supplementary Table 7).

Gene-based testing largely reinforced the established single-variant architecture by concentrating association evidence into a limited number of genes within known loci (e.g., *PHACTR1* on chr 6p24 and *LRP1* on chr 12q13; Supplementary Table 7). Overall, 34 of the 46 Bonferroni-significant genes mapped within 500 kb of at least one genome-wide significant SCAD SNP identified in the SCAD-only or SCAD-MTAG analyses, indicating broad concordance between variant-level and gene-level discovery in these regions (Supplementary Table 7).

Importantly, LDAK-GBAT also revealed 12 genes that remained significant after Bonferroni correction across all tested genes (P < 2.79×10⁻⁶), while lying outside any locus defined by genome-wide significant SNPs in the SCAD-only or MTAG analyses. These represent additional loci that are detectable only when aggregating SNP effects at the gene level, and therefore demonstrated the added power of this gene-based association analysis. The top-most significant genes included the ABO, alpha 1-3-N-acetylgalactosaminyltransferase and alpha 1-3-galactosyltransferase gene (*ABO)*, which determines blood group and is a major regulator of von Willebrand factor and factor VIII levels ^43–45^, and the oestrogen receptor β (ERβ) gene, (*ESR2*), reported to mediate oestradiol-dependent inhibition of vascular smooth muscle cell proliferation ^46^. These findings highlight the potential contribution of coagulation and hormonal pathways to SCAD susceptibility beyond the established risk loci.

### SCAD risk variants are enriched in regulatory elements active in coronary-artery smooth-muscle cells

For each of the 40 loci significant from MTAG, we defined candidate functional variants as the union of the SNPs belonging to the 95% credible-set and those in high LD (r² > 0.8) with the lead SNP in European-ancestry samples from the 1000 Genomes Project (Supplementary Table 4). We then tested whether these variants were enriched in cell-type–resolved open chromatin from coronary-artery and the histone mark for active regulatory regions (H3K27ac) in any tissue from ENCODE (Figure 2).

**Figure 2.**
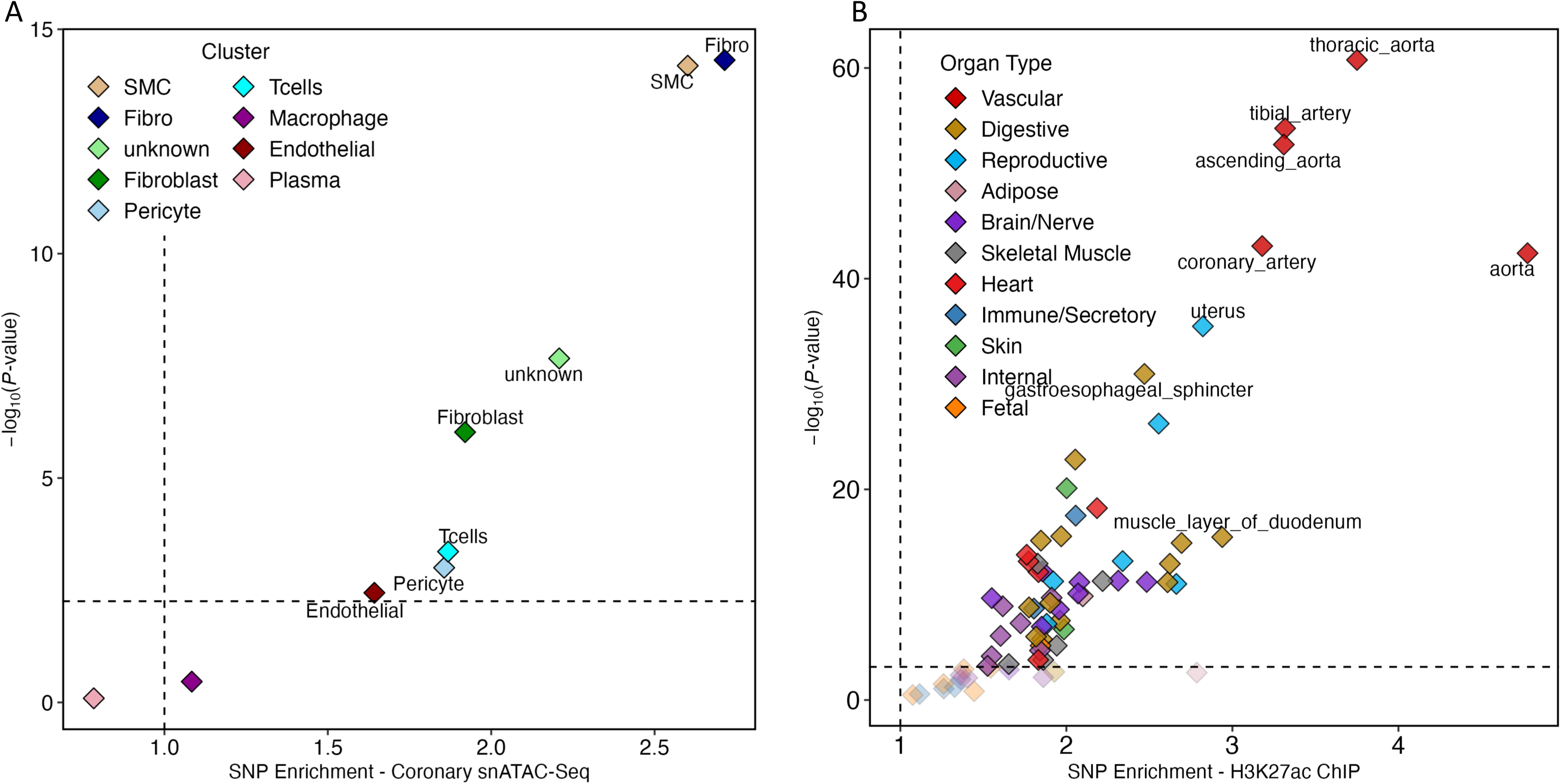
SCAD risk variants are enriched in coronary-artery smooth-muscle regulatory elements. (**A**) Enrichment of SCAD-associated variants in chromatin accessible in single-nucleus ATAC–seq clusters from human coronary artery. Each point represents a cell cluster, coloured by cell type. (B) Enrichment of the same set of variants in H3K27ac ChIP–seq peaks from ENCODE tissues. Each point represents one tissue sample, coloured by organ group. In both panels, the x-axis shows the fold enrichment of candidate SCAD variants in the corresponding regulatory peaks, and the y-axis shows the –log₁₀ P value for the enrichment. Vertical and horizontal dashed lines indicate no enrichment (fold = 1) and the nominal significance threshold, respectively.

We found that SCAD variants were most significantly enriched in peaks specific to smooth-muscle and fibroblast-like clusters defined in coronary-artery snATAC–seq data, and showed the largest fold enrichment, compared with matched control SNPs (Figure 2A). Enrichments in endothelial, immune and other non-contractile clusters were generally weaker and close to the null expectation. At the tissue-sample level, SCAD variants were preferentially enriched in H3K27ac-marked active regulatory regions from vascular samples, including coronary artery and aortic tissues, whereas most non-vascular samples showed little to no enrichment (Figure 2B). Taken together, these findings indicate that SCAD risk variants are preferentially positioned in regulatory elements active in coronary-artery smooth muscle and stromal compartments, rather than in broadly active regulatory elements across unrelated tissues.

### Cis-eQTLs and colocalization support expression-mediated effects at SCAD loci

To investigate whether SCAD risk alleles act through gene expression, we also intersected candidate variants from all the 40 loci with cis-eQTLs in ten vascular and cardiometabolic tissues (GTEx, v10 ^40^). Across all variants within the locus windows, we identified 390 significant cis-eQTL gene–tissue pairs involving 136 genes (Supplementary Table 8). The largest numbers were observed in tibial artery (64 pairs), tibial nerve (60), cultured fibroblasts (55) and aorta (54). These account for about 60% of all pairs.

We performed colocalization between SCAD association signals at SCAD MTAG loci and GTEx v10 cis-eQTLs that are restricted to significant cis-eQTL gene–tissue pairs in GTEx (Methods). We identified 99 gene–tissue pairs with strong evidence for a shared causal variant (posterior probability for H4 ≥ 0.8), corresponding to 47 unique genes (Supplementary Table 8). These pairs were detected across all ten tissues but were most frequent in arterial samples: 59 of 99 pairs (59%) mapped to aorta, tibial artery and coronary artery, followed by tibial nerve (12 pairs), cultured fibroblasts (8) and cardiac tissues (12 pairs in atrial appendage and left ventricle combined; Supplementary Table 8, Supplementary Figure 3).

For the 24 newly identified loci, Figure 3 summarizes 54 colocalizing gene–tissue pairs involving 27 genes. The broadest multi-tissue signals were observed for *ZNF638* and *GGCX* (five tissues each), followed by *TNFSF12* (four tissues), and *FRK*, *NT5DC1*, *SPON1* and *SLC24A3* (three tissues each). Across these novel loci, the direction of the eQTL effect was in many cases consistent across tissues. At several genes, the SCAD risk allele was associated with lower expression, including *GGCX*, *FRK*, *SPON1*, *FGF9*, *TGFB3*, *BCAR1* and *SLC24A3*. In contrast, at *SNX13*, *EHBP1L1*, *ADAMTS8* and *TNFSF12*/*TNFSF13*, the risk allele was associated with higher expression. *ZNF638* was an exception, showing tissue-dependent direction (Figure 3A).

**Figure 3.**
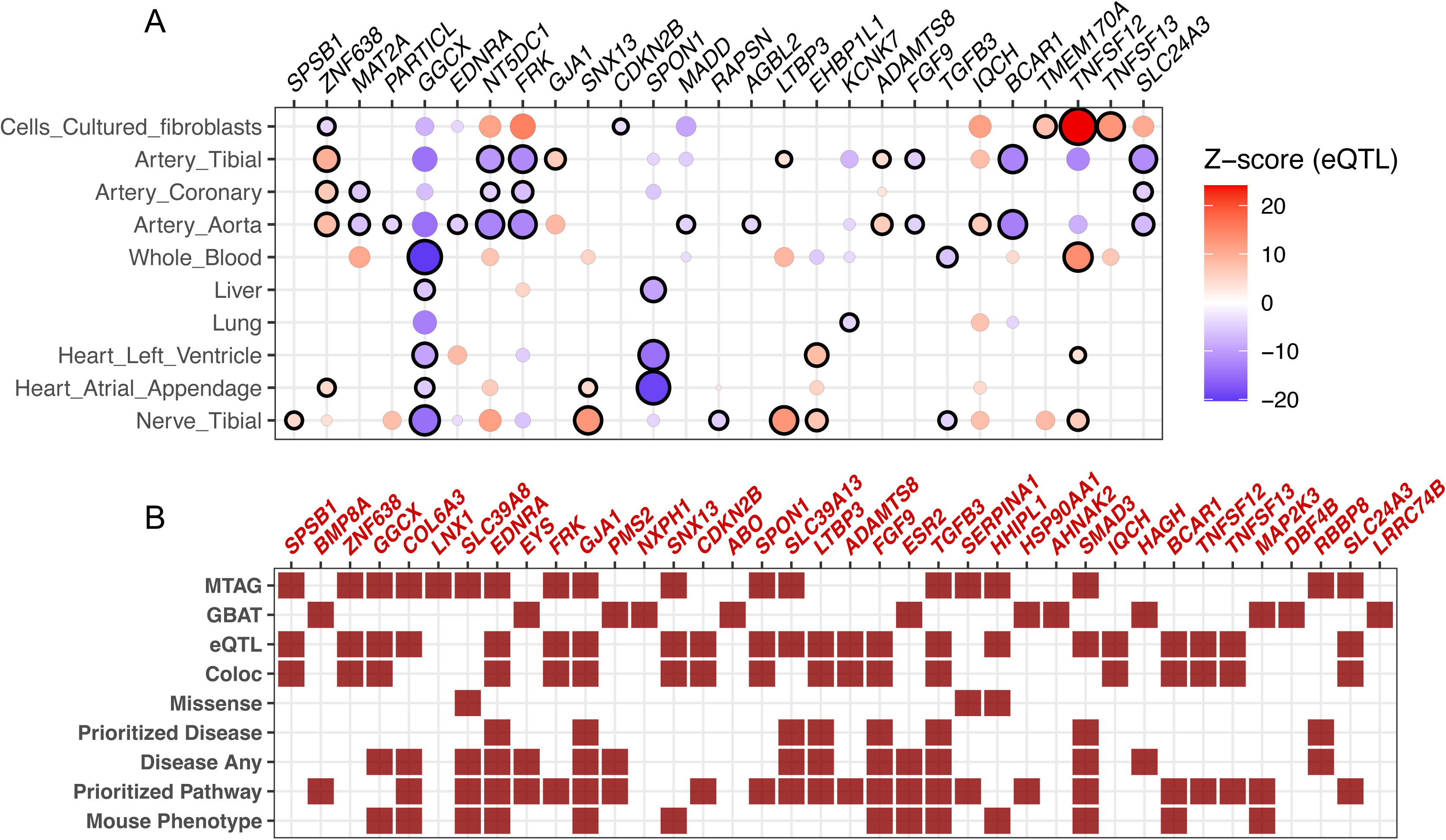
eQTL colocalization and integrated evidence for prioritized genes. **A**, Dot plot showing gene–tissue pairs with evidence of colocalization between SCAD MTAG association signals and GTEx v10 cis-eQTLs at loci that are newly associated with SCAD. Columns correspond to genes and rows to tissues. For each gene, the plotted variant is the top candidate SNP, defined as the variant that minimises the product of the SCAD MTAG and eQTL P-values for that gene. Each circle represents the eQTL association of this variant with gene expression in a given tissue; circle colour indicates the eQTL Z-score aligned to the SCAD risk-increasing allele (red, higher expression with the risk allele; blue, lower expression), as shown on the colour scale. Circle size reflects the strength of the eQTL signal. Circles outlined in black mark gene–tissue pairs with strong support for a shared causal variant between SCAD and the eQTL (colocalization posterior probability for H4 ≥ 0.8). Only genes with colocalization in at least one tissue are displayed. **B,** Tile plot summarizing the lines of evidence supporting 38 genes prioritized at SCAD loci that were not genome-wide significant in the SCAD-only GWAS. Each column represents one prioritized gene and each row a source of evidence. Filled red tiles indicate that the gene satisfies the corresponding criterion, blank tiles indicate absence of evidence. The upper five rows correspond to annotations directly used for prioritization: “MTAG” denotes the nearest gene to the MTAG lead SNP; “GBAT” indicates significance in LDAK-GBAT gene-based testing; “eQTL” indicates at least one significant cis-eQTL in GTEx v10 involving a candidate variant; “Coloc” marks loci with strong colocalization between SCAD MTAG and cis-eQTL signals (posterior probability for a shared causal variant H4 ≥ 0.8); and “missense” indicates presence of a candidate missense variant among SCAD candidate variants. The lower four rows summarize downstream annotation: “Prioritized disease” indicates membership of a disease term significantly enriched among prioritized genes; “Disease any” indicates any ClinVar disease association; “Prioritized pathway” indicates membership of an enriched biological pathway; and “Mouse phenotype” indicates a cardiovascular phenotype in Mouse Genome Informatics.

These results support a model in which, at many SCAD loci, the same variants influence both disease risk and local gene expression across the ten tissues examined, with signals observed most frequently in arterial tissues and additional colocalizations in tibial nerve, cultured fibroblasts and heart.

### Integrated functional evidence refines SCAD candidate genes

To integrate association and functional evidence, we prioritized candidate genes at each locus by combining proximity to lead SCAD variants, LDAK-GBAT gene-based association, and coding and regulatory annotations, including GTEx cis-eQTL support, colocalization with SCAD MTAG signals and the presence of candidate missense variants (Figure 3B, Supplementary Figure 5). We found that newly identified SCAD loci involved both candidate missense variants and proximal or distal regulatory variants. Candidate missense variants were observed at four genes (Supplementary Table 9). In particular, at chr 14q32.13, the SCAD association signal included a low-frequency variation tagged by rs28929474, a SERPINA1 missense variant with prior clinical annotation as likely pathogenic and known to underlie α1-antitrypsin deficiency ^47^, and is also associated with circulating protein levels including sex hormone-binding globulin ^48^.

A majority of MTAG/LOO novel loci involved tissue-specific regulatory evidence, including *GGCX*, *EDNRA*, *LTBP3*, *SPON1*, *FRK*, *SLC24A3*, *SLC39A13* and *TNFSF12*. For example, at chr4q31, the association peak upstream of *EDNRA* mapped to a compact proximal regulatory element active in coronary-artery smooth-muscle and pericyte chromatin, with concordant arterial cis-eQTL colocalization for *EDNRA*. The rs6841581 risk allele is predicted to create a motif for HEY1/2 transcriptional repressors, which act downstream of NOTCH signalling ^49^ (Supplementary Figure 6).

In some cases, cis-eQTL colocalization occurred outside of typical arterial tissues, suggesting systemic mechanisms to play a role in SCAD. For example, at chr2p13.3, the lead association maps upstream of *GGCX*; the SCAD signal colocalizing with *GGCX* cis-eQTLs across multiple tissues, including liver, and multi-trait fine-mapping prioritized candidate variants in the *GGCX* 3′UTR, consistent with post-transcriptional regulation (Figure 3, Supplementary Figure 7).

### Enrichment of SCAD prioritized genes in functional pathways

Altogether, combined annotations yielded 56 prioritized genes across the 40 SCAD loci, including 38 genes at loci newly implicated by MTAG panel or LDAK-GBAT and 18 at previously reported SCAD loci (Supplementary Tables 10–11). Overall, 39 of 56 prioritized genes showed regulatory evidence from cis-eQTL analysis, colocalization, or both; of these, 29 showed both cis-eQTL support and colocalization, 10 showed cis-eQTL support alone, and 17 showed neither. Among the 38 genes at newly implicated loci, 22 showed regulatory evidence from cis-eQTL analysis, colocalization, or both, including 18 with both cis-eQTL support and colocalization (H4 ≥ 0.8 in at least one tissue) and 4 with cis-eQTL support alone; 16 showed neither. In addition, 12 of the 38 novel-locus genes reached Bonferroni significance in LDAK-GBAT.

Among top significantly enriched pathways involving the 56 prioritized genes we cite three groups of GO terms: extracellular-matrix biology (“collagen-containing extracellular matrix”, “extracellular matrix, structural constituent” and “extracellular matrix organization”), bone biology (“bone mineralization” and “regulation of bone mineralization”, supported by a connected subnetwork including *ECM1*, *ATP2B1*, *SLC24A3*, *GJA1* and TGF-β-linked genes such as *LTBP3* and *SMAD3)*, and TGF-β signalling (“cellular response to transforming growth factor beta stimulus”, “cellular surface receptor protein serine threonine kinase signalling pathway”) (Figure 4A). Consistently, SCAD genes were enriched among disease terms related to vascular and heritable connective tissue disorder, including familial thoracic aortic aneurysm and aortic dissections, reflecting overlap with established arteriopathy genes (e.g., *TGFB3*, *FBN1*, *LTBP3*, *COL6A3*, *GJA1*, Figure 4B).

**Figure 4.**
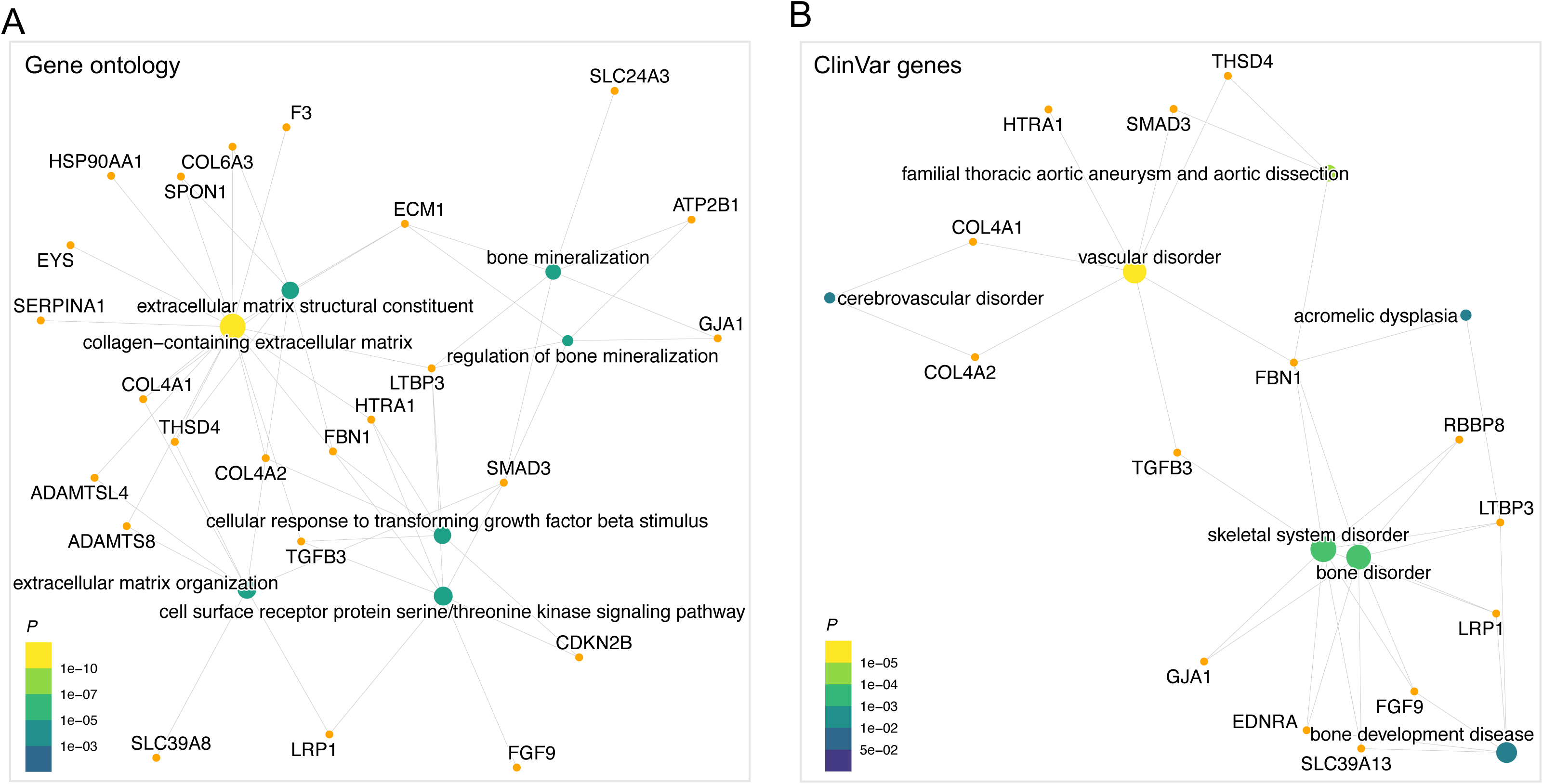
Gene Ontology and Disease term enrichment of genes prioritized at SCAD loci. Concept network plots linking the top enriched Gene Ontology terms (A) and top ClinVar annotations (B) among the 56 genes prioritized from MTAG and gene-based analyses (text-labelled nodes, colours-scale indicates enrichment P-value) to their member genes (gene-labelled nodes). Edges indicate gene membership in each term and highlight genes shared across multiple enriched pathways.

## DISCUSSION

This study leverages genetic overlap of SCAD with several vascular traits in a multi-trait GWAS and gene-level association framework to substantially boost our currently limited knowledge, resulting from the unavailability of large samples. Through an integrated and multi-resourced approach we successfully (*i*) broaden the SCAD genetic catalogue to 40 risk loci and 56 candidate genes, (*ii*) strengthen the evidence for the regulatory role of risk variants in the arterial wall, mainly in coronary-artery smooth-muscle cells and fibroblasts, and (*iii*) connect SCAD risk to interpretable arterial-wall biological processes like TGF-β signalling and bone mineralization and osteogenic programs, while confirming further the central role of extracellular matrix biology and providing additional genetic evidence for blood coagulation regulation.

From a genetic-architecture standpoint, the multi-trait framework suggests that a substantial fraction of SCAD susceptibility was already present as directionally consistent but sub-genomic threshold signals in the original meta-GWAS. Joint modelling with genetically related vascular traits converted many of these signals into firmly associated loci while preserving essentially all established associations, with a significant proportion showing consistent direction in an independent case control study. The largest gains came from aneurysmal and dissection phenotypes of the aorta, and cerebral vascular traits. This pattern is consistent with substantial pleiotropy across arterial diseases, indicating that many of the loci identified here are not unique to SCAD but lie within a broader shared vascular genetic architecture. In contrast, atherosclerotic coronary artery disease contributed less and was often directionally discordant, consistent with the already established negative genome-wide genetic correlation ^13^. Leave-one-out MTAG analyses further highlighted that cross-trait consistency is locus dependent, with as many as ten loci reaching genome-wide significance only after removal of a specific auxiliary trait. At several loci, locus-specific divergence between SCAD and CAD was associated with weaker SCAD signal detection in the full model, whereas the association became more pronounced when CAD was excluded (e.g., *F3*, *CDKN2B* and *TGFB3*). At another subset of loci, divergence between SCAD and TAAD reduced sensitivity until TAAD was removed (e.g. loci near *SLC24A3* and *GJA1*). Elsewhere, excluding a given auxiliary trait produced only modest gains, suggesting that the trait was less informative for SCAD signal detection at those specific loci rather than globally uninformative. These observations support the view that the vascular traits included in our framework capture broadly shared biology with SCAD, while their contribution to discovery can vary across loci. This further illustrates the value of locus-aware multi-trait refinement, even when effect directions differ across traits.

Our gene-based testing adds a complementary perspective to boosted discovery of SCAD relevant genes. While this method sharpens effector gene assignment at known loci (e.g., *PHACTR1*, *LRP1*), it also reveals additional genes that are undetected at the single-variant level but harbour a significant burden of associated variants when aggregated. We note that the gene-based analysis provided complementary associated genes, with limited overlap with prioritized genes at MTAG loci. While the aggregation feature of this method enhanced the detection power of modest signals located within genic definition as available in Ensembl v105 RefSeq annotation, we acknowledge the limitations of this method to capture genetic signals located in intergenic regions as in the case of *COL6A3* and *EDNRA* loci.

Among associated genes, we highlight the oestrogen receptor *ESR2*, which was previously linked to the regulation of salt-sensitive blood pressure in women^50^, and the blood type locus *ABO*, which is central to several haemostatic traits ^43,51,52^. Although these genes were detected only by gene-based testing, variants contributing to their gene-based signals showed regulatory annotations and expression-based support consistent with both the original SCAD GWAS^13^ and the multi-trait loci reported here. Thus, the aggregation of many small effects seems to be an important component of SCAD genetic risk, which supports further its highly polygenic architecture.

Pathway and disease enrichment analyses involving the updated list of prioritized genes reinforce extracellular-matrix (ECM) structure and organization as central themes in SCAD pathogenesis, consistent with prior evidence implicating ECM and collagen-fiber abnormalities in SCAD and related arteriopathies ^13,53^. Among newly prioritized genes in this subset of genes, collagen 6 subunit gene *COL6A3* is an established cause of inherited small-vessel disease, aortopathies, and connective-tissue disorders ^54–58^. One interesting result is that several loci also implicate TGF-β–related growth-factor signalling, involving *LTBP3*, *SMAD3* and *TGFB3*, which reinforces TGF-β pathway-involvement, consistent with its established role in Mendelian aortopathies ^59–65^. Our multi-trait design, leveraging shared architecture with aneurysmal phenotypes has improved sensitivity to cover these highly relevant pathways to arterial dissection.

We found that several bone mineralisation-related terms and diseases as significantly enriched for SCAD genes. This is driven both by genes common to the TGF-β and bone morphogenetic protein (BMP) pathways, such as *LTBP3* and *SMAD3*, but also genes involved in calcium handling such as *ATP2B1* and *SLC24A3*, which encode transmembrane channels exporting calcium from the cytosol to the extracellular space. *GJA1* encodes a gap-junction protein, connexin 43, which was shown to play a role in osteoblast differentiation in mice and humans ^66,67^. Osteogenic pathways and calcium trafficking play a major role in vascular calcification, a pathological process associated with vascular aging and atherosclerosis driven by vascular smooth muscle phenotypic changes ^68^. Interestingly, the oestrogen receptor-β, encoded by *ESR2* gene, was also linked to vascular calcification in mice ^69^. Further studies will be required to determine whether SMC phenotypic changes at stake in vascular calcification could play a role in coronary dissection.

Our results further support a haemostatic component to SCAD susceptibility, mainly through prioritization of *ABO* and the gamma-glutamyl carboxylase gene *GGCX*. In addition to blood groups, *ABO* influences plasma levels of von Willebrand factor and factor VIII ^43–45^ and has been repeatedly linked to thrombotic cardiovascular outcomes, including acute coronary syndromes and ischemic stroke ^51,52,70–74^. These data motivate targeted follow-up to map the SCAD risk haplotype onto ABO blood-group–tagging variants and to test whether the association aligns with established prothrombotic profiles. Our work links for the first time SCAD risk to *GGCX*, which encodes the enzyme responsible for vitamin K–dependent γ-carboxylation. GGCX activates multiple vitamin K–dependent coagulation factors, in addition to components of the arterial-wall biology such as matrix Gla protein (MGP), a key inhibitor of osteogenic BMP signalling and vascular calcification ^75,76^. In our colocalization results, the SCAD risk–increasing allele was associated with lower *GGCX* expression in arterial tissues, consistent with reduced γ-carboxylation capacity ^75,77^. Pharmacologic inhibition of the same pathway by vitamin K antagonists (VKA) is associated with increased vascular and valvular calcification in humans, supporting that reduced γ-carboxylation can modify vessel-wall properties ^78–81^. Together, these observations motivate targeted follow-up of vitamin K–related exposures (e.g. vitamin K status and VKA therapy) as potential modifiers of arterial-wall remodelling in SCAD, and the propensity for intramural hematoma propagation once dissection is initiated. The net direction and magnitude of such effects on SCAD risk, notably in case of recurrent SCAD, needs further clinical investigation.

Our findings also highlight vasoactive tone and mechano-transduction genes as potential contributors to SCAD risk. *EDNRA* encodes the endothelin receptor ET-A, the major receptor to endothelin-1 (ET-1) in vascular SMCs. The tight regulation of vasoconstriction and vascular smooth-muscle tone by endothelium-derived ET-1 relies on the balance between the two receptors ET-A, a direct inducer of intracellular calcium release and vascular SMC contraction, and ET-B, which trigger the release of nitric oxide from the endothelium, counteracting ET-A dependent contraction^82^. Smooth-muscle–specific *Ednra* knockout in mice leads to arterial tortuosity, altered arterial patterning and complete impairment of endothelin-1–mediated vasoconstriction^83^. ET-1 signalling was involved in pulmonary arterial hypertension (PAH), highlighting its impact on the modulation of vascular SMC phenotype and proliferation potential beyond short-term contractile effect^84^. A wide range of ET-1 receptor specific antagonists have been developed and tested in the context of human diseases, notably PAH^85^, making this gene a promising drug target in the context of SCAD. Additional genes identified as target genes at SCAD loci play a role in vascular SMC contraction, such as *BCAR1*, which encodes the adaptor protein p130Cas^86,87^, and genes encoding calcium channels *SLC24A3* and *ATP2B1*^88,89^. These observations support altered sensing and transduction of hemodynamic forces in coronary arteries as a genetically supported mechanism contributing to SCAD.

A key unresolved question is why SCAD risk manifests predominantly in women, particularly around reproductive transitions. While our analyses do not directly test sex-specific mechanisms, some of them lead to testable hypotheses linking arterial-wall regulatory biology to female hormonal and hemodynamic milieus. The potential role of oestrogen receptor-β, encoded by *ESR2*, in SCAD could represent a first direct link with female hormones, which have long been suspected to play a triggering role in postpartum SCAD ^90^. On the other hand, *ABO*-related variation in von Willebrand factor and factor VIII ^43,51,52^, and pregnancy-associated thrombotic risk differences by blood group ^91–93^, motivate evaluation of haemostatic pathways in pregnancy-associated and postpartum SCAD. Endothelin signalling is modulated by oestrogen and other sex hormones ^94–97^, raising the possibility that ET-A-mediated vasoreactivity could show context-dependent effects across hormonal states. Future work integrating genetic risk with reproductive history, hormonal profiling and longitudinal vascular imaging could help test these hypotheses in well-phenotyped cohorts.

Our genetic results and their biological inferences should be interpreted in light of several limitations. First, MTAG assumes partially shared genetic architecture across traits and can yield biased estimates when this assumption is strongly violated ^17^. To mitigate this risk, we restricted analyses to vascular phenotypes with clear clinical and genetic overlap with SCAD and verified concordant effect directions between MTAG and the SCAD-only GWAS for lead signals, although some loci may still reflect complex pleiotropy or residual confounding effects. Secondly, while our independent look-up in an independent case-control dataset provides supportive direction-of-effect evidence, it is only modestly powered for single-variant replication. Future validation in larger studies is needed, including in more ethnically diverse backgrounds, where we lack genetic data for SCAD. Indeed, the available GWAS were predominantly of European ancestry, limiting generalizability and motivating future multi-ancestry studies. Finally, we prioritized genes using a systematic annotation with publicly available genomic and epigenomic resources, which do not capture the full, context-specific regulatory mechanisms in coronary arteries and do not necessarily reflect the complexity of the regulatory mechanisms at stake at SCAD loci. Consequently, assigning association signals to effector genes remains uncertain at some loci, and multiple genes or multiple causal variants may contribute to a given association signal.

In summary, the present work provides an updated and integrated genetic and functional framework for SCAD. Our findings place SCAD on a continuum with inherited arteriopathies and connective-tissue disorders, while distinguishing it from atherosclerotic CAD. They also provide a substantial list of highly relevant genes and pathways for experimental follow-up in vascular cells and model systems. Future work integrating these genetic insights with detailed phenotyping, imaging and environmental exposures should help clarify how polygenic risk interacts with hormonal and hemodynamic triggers to precipitate coronary artery dissection, and may ultimately inform risk stratification and target key strategy-preventions in early middle-aged women, the main target population in SCAD.

## FUNDING

This study was supported by “Fondation pour la Recherche Médicale” individual fellowship to TEB (ARF202309017669), Agence Nationale de la Recherche “jeune chercheur” to AG (ANR-23-CE17-0004), National Health and Medical Research Council postgraduate scholarship (2039699) with co-funding from a National Heart Foundation PhD scholarship (108543) to IT, an NSW Health Early Mid-Career Cardiovascular Grant and a NHMRC Investigator Grant (2018360) to EG, an NHMRC Grants (APP1161200, APP2010203) and an NSW Health Cardiovascular Senior Scientist Grant to RMG, funds from SCAD Research Inc., Australia to RMG, EG and IT, European Research Council Consolidator Grant (ID 101088901, ClassifyDiseases) to DS, European Research Council Starting Grant (ID: 716628, ROSALIND) to NBN and “Fondation Lefoulon Delalande” as part of an ICRPA funding scheme (IA/F/24/275131) to NBN.

## Data Availability

Instructions for running LDAK-GBAT are available on the LDAK website, while sample code for the analyses in this paper are on the GitHub page of TEB. We have access to UKBB data via application 21,432. We applied for and downloaded MVP summary statistics from the dbGaP website. Summary statistics of MTAG results will be submitted to GWAS catalog upon article publication.

## ACKNOWLEDGEMENTS

We thank the participants and investigators of the SCAD studies contributing to the discovery analyses and the Victor Chang Cardiac Research Institute Arteriopathies & SCAD Cohort (VASC). We also thank the original investigators who shared intracranial aneurysm and cervical artery dissection summary statistics. We acknowledge UK Biobank, GTEx, ENCODE, and the Million Veteran Program/dbGaP for access to data and resources used in this study.

## AUTHOR CONTRIBUTIONS

Study design/conception: TEB and NBN. Data analyses: TEB, AG, IT, EG, DS. Writing and editing the manuscript: TEB, AG and NBN. All authors critically reviewed and edited the manuscript and approved the final version.

## DISCLOSURE OF INTEREST

The authors declare no competing interests.

## WEB RESOURCES

MTAG, https://github.com/JonJala/mtag

LocusZoom, http://locuszoom.org/

LDAK-GBAT, https://www.ldak.org

GREGOR, https://csg.sph.umich.edu/GREGOR/

Coloc, https://chr1swallace.github.io/coloc/

FUSION, http://gusevlab.org/projects/fusion/

UK Biobank, https://www.ukbiobank.ac.uk/

Million Veteran Program summary statistics, https://www.ncbi.nlm.nih.gov/projects/gap/cgi-bin/study.cgi?study_id=phs001672.v3.p1

ENCODE, https://www.encodeproject.org/

GTEx, https://www.gtexportal.org/home/downloads/adult-gtex/qtl

## APPENDICES

None

## Notes

### Competing Interest Statement

The authors have declared no competing interest.

### Funding Statement

This study was supported by Fondation pour la Recherche Medicale individual fellowship to TEB (ARF202309017669), Agence Nationale de la Recherche jeune chercheur to AG (ANR-23-CE17-0004), National Health and Medical Research Council postgraduate scholarship (2039699) with co-funding from a National Heart Foundation PhD scholarship (108543) to IT, an NSW Health Early Mid-Career Cardiovascular Grant and a NHMRC Investigator Grant (2018360) to EG, an NHMRC Grants (APP1161200, APP2010203) and an NSW Health Cardiovascular Senior Scientist Grant to RMG, funds from SCAD Research Inc., Australia to RMG, EG and IT, European Research Council Consolidator Grant (ID 101088901, ClassifyDiseases) to DS, European Research Council Starting Grant (ID: 716628, ROSALIND) to NBN and Fondation Lefoulon Delalande as part of an ICRPA funding scheme (IA/F/24/275131) to NBN.

### Author Declarations

Source of summary statistics https://ftp.ebi.ac.uk/pub/databases/gwas/summary_statistics/GCST90245001-GCST90246000/GCST90245878/ http://ftp.ebi.ac.uk/pub/databases/gwas/summary_statistics/GCST90026001-GCST90027000/GCST90026612 http://www.nealelab.is/uk-biobank https://cardiogramplusc4d.org/data-downloads/ https://csg.sph.umich.edu/willer/public/AAAgen2023/phs001672.v10.p1 (https://www.ncbi.nlm.nih.gov/projects/gap/cgi-bin/study.cgi?study_id=phs001672.v10.p1

