## Supplementary figures for "Multi-trait and Gene-Based Analyses Identify Genetic Variants Associated with Spontaneous Coronary Artery Dissection"

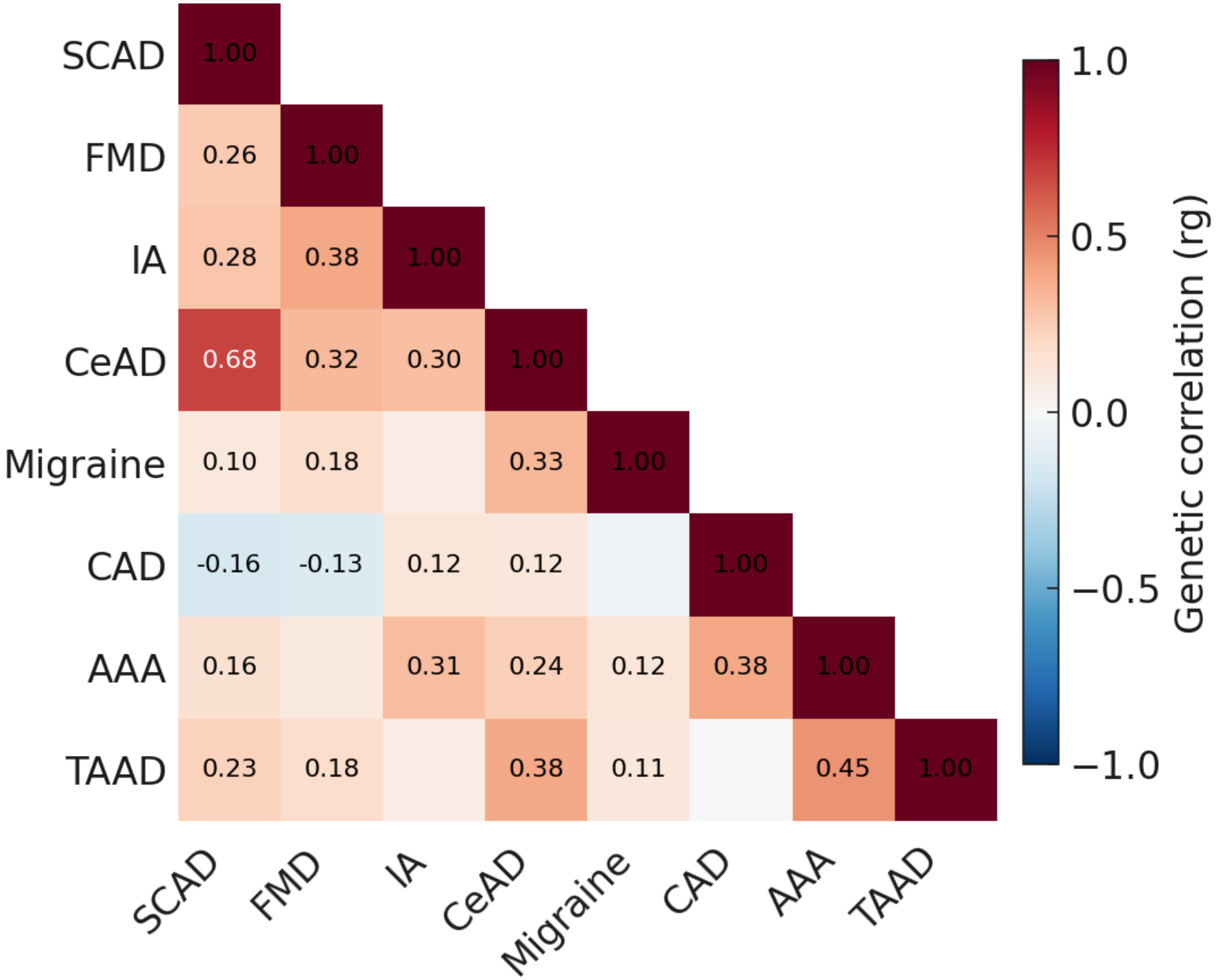


**Supplementary Figure 1. Genome-wide genetic correlation matrix among SCAD and auxiliary traits used for MTAG.**
Pairwise genome-wide genetic correlations (rg) were estimated between spontaneous coronary artery dissection (SCAD) and the seven auxiliary traits using linkage disequilibrium score regression (LDSC) on GWAS summary statistics. Auxiliary traits were fibromuscular dysplasia (FMD), intracranial aneurysm (IA), cervical artery dissection (CeAD), migraine, coronary artery disease (CAD), abdominal aortic aneurysm (AAA), and thoracic aortic aneurysm/dissection (TAAD). Values shown in each cell correspond to the LDSC point estimate of rg. Colour intensity reflects the magnitude and sign of rg (red, positive correlation; blue, negative correlation).

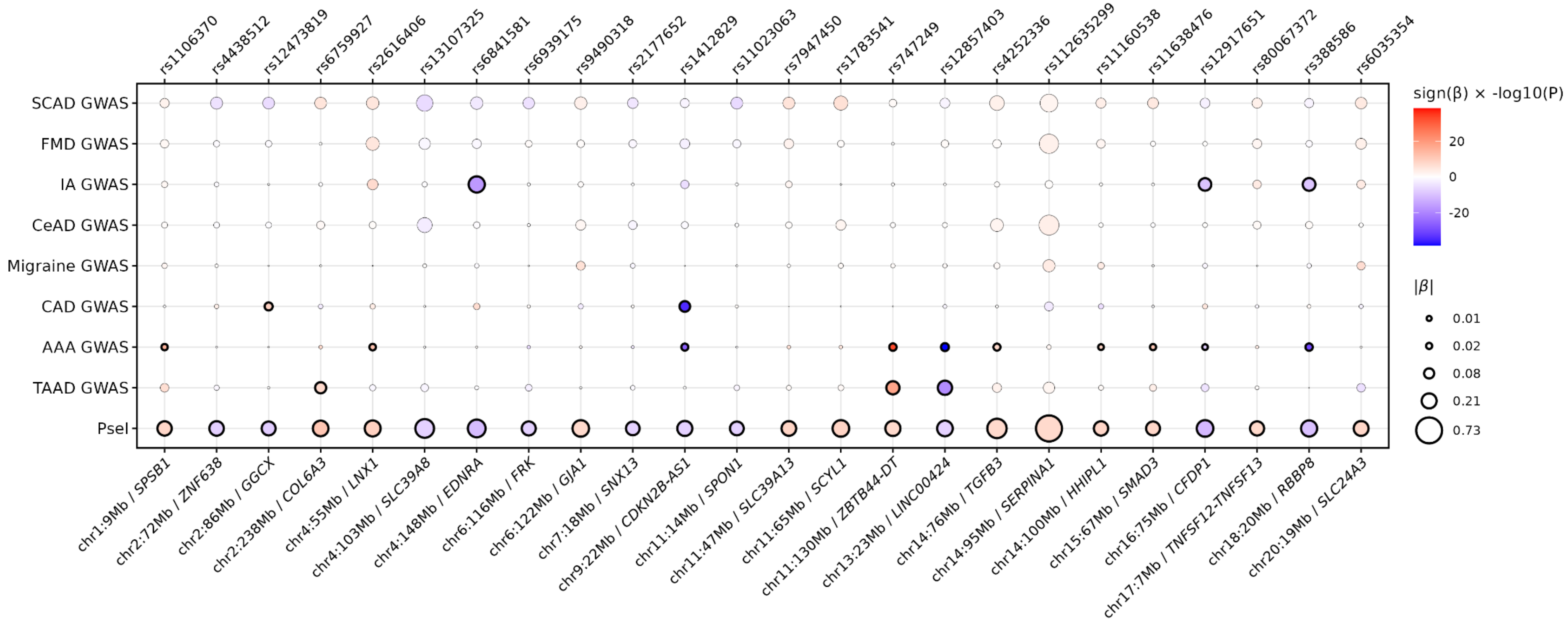


**Supplementary Figure 2. Sharing of multi-trait SCAD signals with the contributing traits.**

Dot plot summarizing association evidence for the 24 lead variants at loci that were novel for spontaneous coronary artery dissection (SCAD) in the multi-trait discovery analysis. Each column corresponds to one lead single-nucleotide polymorphism (SNP) (rsID shown on the top x-axis; genomic position and closest gene shown on the bottom x-axis). Rows show results from the SCAD single-trait genome-wide association study (GWAS), the seven auxiliary single-trait GWASs (fibromuscular dysplasia (FMD), intracranial aneurysm (IA), cervical artery dissection (CeAD), migraine, coronary artery disease (CAD), abdominal aortic aneurysm (AAA), and thoracic aortic aneurysm and dissection (TAAD)), the primary 8-trait multi-trait analysis of GWAS (MTAG) analysis (SCAD plus all seven auxiliary traits), and the best-performing MTAG configuration for that SNP ("Best MTAG model", defined as the most significant result across the primary MTAG analysis and the seven leave-one-out MTAG runs). For each SNP–analysis cell, circle area is proportional to the absolute effect estimate |β| for the effect allele (alleles harmonized across traits to the SCAD effect allele). Circle colour encodes signed strength of association, sign(β)×$-\log_{10}$(P) (red, positive; blue, negative; white, near zero). All circles are shown with a thin outline for readability; bold black outlines indicate genome-wide significance within the corresponding analysis (two-sided $P<5\times10^{-8}$). The plot provides an at-a-glance view of (i) how apparent each signal is in the single-trait SCAD GWAS, (ii) concordant or discordant directions of effect across genetically correlated traits, and (iii) the extent to which multi-trait modelling amplifies SCAD association evidence at these loci.

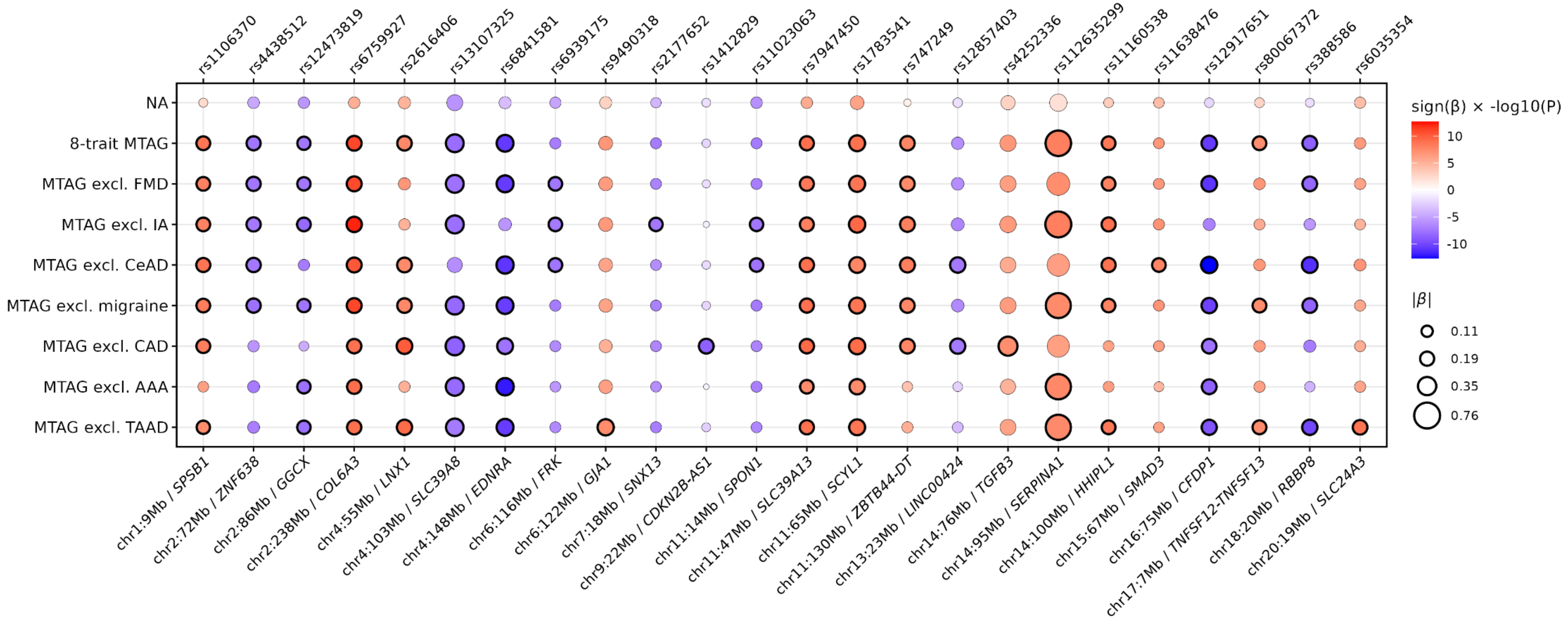


**Supplementary Figure 3. Behaviour of novel SCAD loci across 8-trait MTAG analysis and leave-one-out MTAG configurations.**
Dot plot summarizing SCAD association evidence for the 24 lead variants at loci that were novel for SCAD in the multi-trait discovery analysis, across the SCAD single-trait GWAS, the primary 8-trait MTAG analysis (SCAD plus all seven auxiliary traits), and the seven leave-one-out MTAG runs (each omitting one auxiliary trait). Columns correspond to the same set of lead SNPs as in Supplementary Figure 2 (rsID on the top x-axis; genomic position and closest gene on the bottom x-axis). Rows correspond to the SCAD GWAS, the primary multi-trait MTAG analysis, and the leave-one-out MTAG configurations excluding FMD, IA, CeAD, migraine, CAD, AAA, or TAAD. For each SNP–model cell, circle area is proportional to the absolute SCAD effect estimate |β| (aligned to the SCAD effect allele), and circle colour encodes signed strength of association, sign(β)×$-\log_{10}$(P) (red, positive; blue, negative; white, near zero). All circles are shown with a thin outline for readability; bold black outlines indicate genome-wide significance (two-sided $P<5\times10^{-8}$). This figure highlights how each novel locus behaves across MTAG configurations, illustrating loci whose SCAD association is robust to the removal of individual traits, loci primarily driven by a subset of traits, and loci for which specific LOO analyses yield the strongest SCAD evidence.

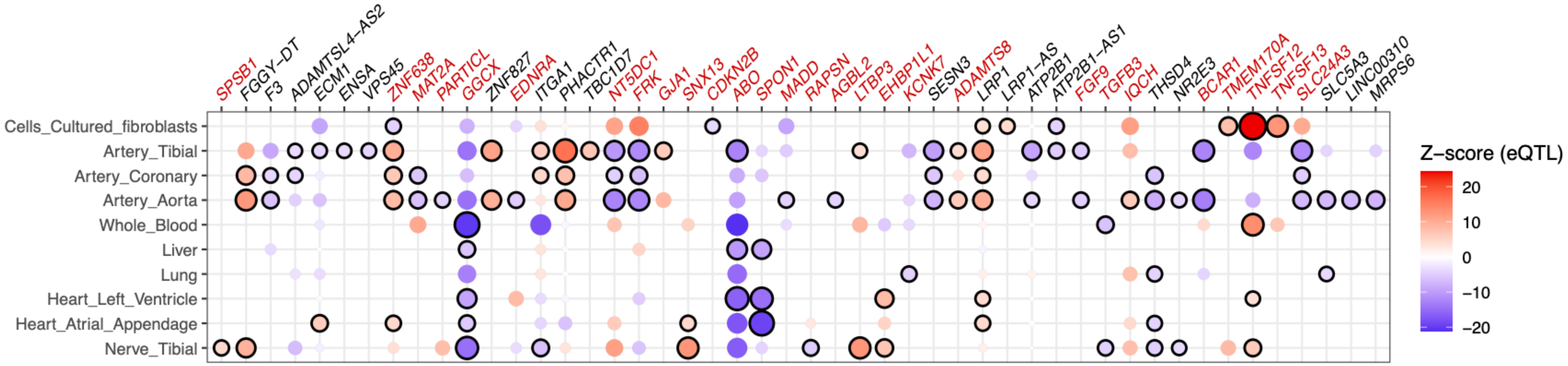


**Supplementary Figure 4. Colocalization between SCAD MTAG signals and cis-eQTLs across all loci.**
Dot plot showing gene–tissue pairs with evidence of colocalization between SCAD MTAG association signals and GTEx v10 cis-eQTLs across all loci. Columns correspond to genes and rows to tissues (cultured fibroblasts, tibial artery, coronary artery, aorta, whole blood, liver, lung, heart left ventricle, heart atrial appendage and tibial nerve). For each gene, the plotted variant is the top candidate SNP, defined as the variant that minimizes the product of the SCAD MTAG and eQTL P-values for that gene. Each circle represents the eQTL association of this variant with gene expression in a given tissue; circle colour indicates the eQTL Z-score aligned to the SCAD risk-increasing allele (red, higher expression with the risk allele; blue, lower expression), as shown on the colour scale, and circle size reflects the strength of the eQTL signal. Circles outlined in black indicate gene–tissue pairs with strong support for a shared causal variant between SCAD and the eQTL (colocalization posterior probability for H4 $\geq 0.8$). Gene names in red denote genes at loci that are newly associated with SCAD in the MTAG analysis. Only genes with colocalization in at least one tissue are shown.

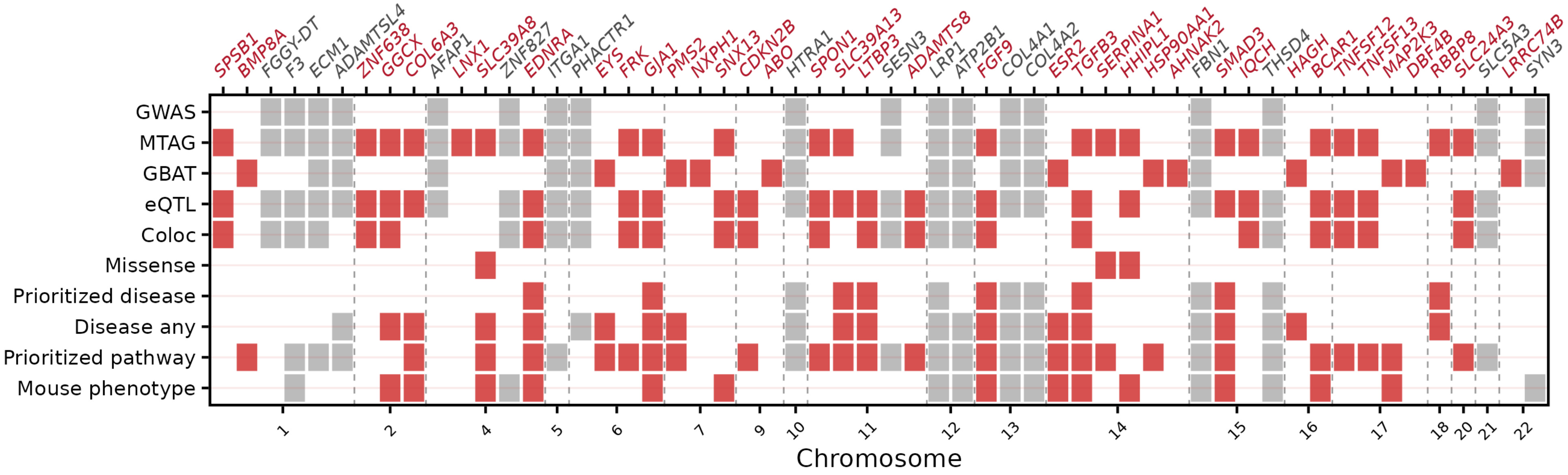


**Supplementary Figure 5. Integrated evidence for all genes prioritized at SCAD loci.**

Tile plot summarizing the lines of evidence supporting 56 genes prioritized across all SCAD loci. Each column corresponds to one gene (x-axis). Gene labels in red indicate loci that are novel for SCAD in the 8 traits MTAG + LOO analysis; labels in grey indicate genes at previously reported SCAD loci and blank tiles indicate absence of evidence. The upper six rows correspond to annotations directly used for prioritization: "GWAS" denotes the nearest gene to the GWAS lead SNP; "MTAG" denotes the nearest gene to the MTAG lead SNP; "GBAT" indicates significance in LDAK-GBAT gene-based testing; "eQTL" indicates at least one significant cis-eQTL in GTEx v10 involving a candidate variant; "Coloc" marks loci with strong colocalization between SCAD MTAG and cis-eQTL signals (posterior probability for a shared causal variant H4 $\geq$ 0.8); and "missense" indicates presence of a candidate missense variant among SCAD candidate variants. The lower four rows summarize downstream annotation: "Prioritized disease" indicates membership of a disease term significantly enriched among prioritized genes; "Disease any" indicates any ClinVar disease association; "Prioritized pathway" indicates membership of an enriched biological pathway; and "Mouse phenotype" indicates a cardiovascular phenotype in Mouse Genome Informatics.

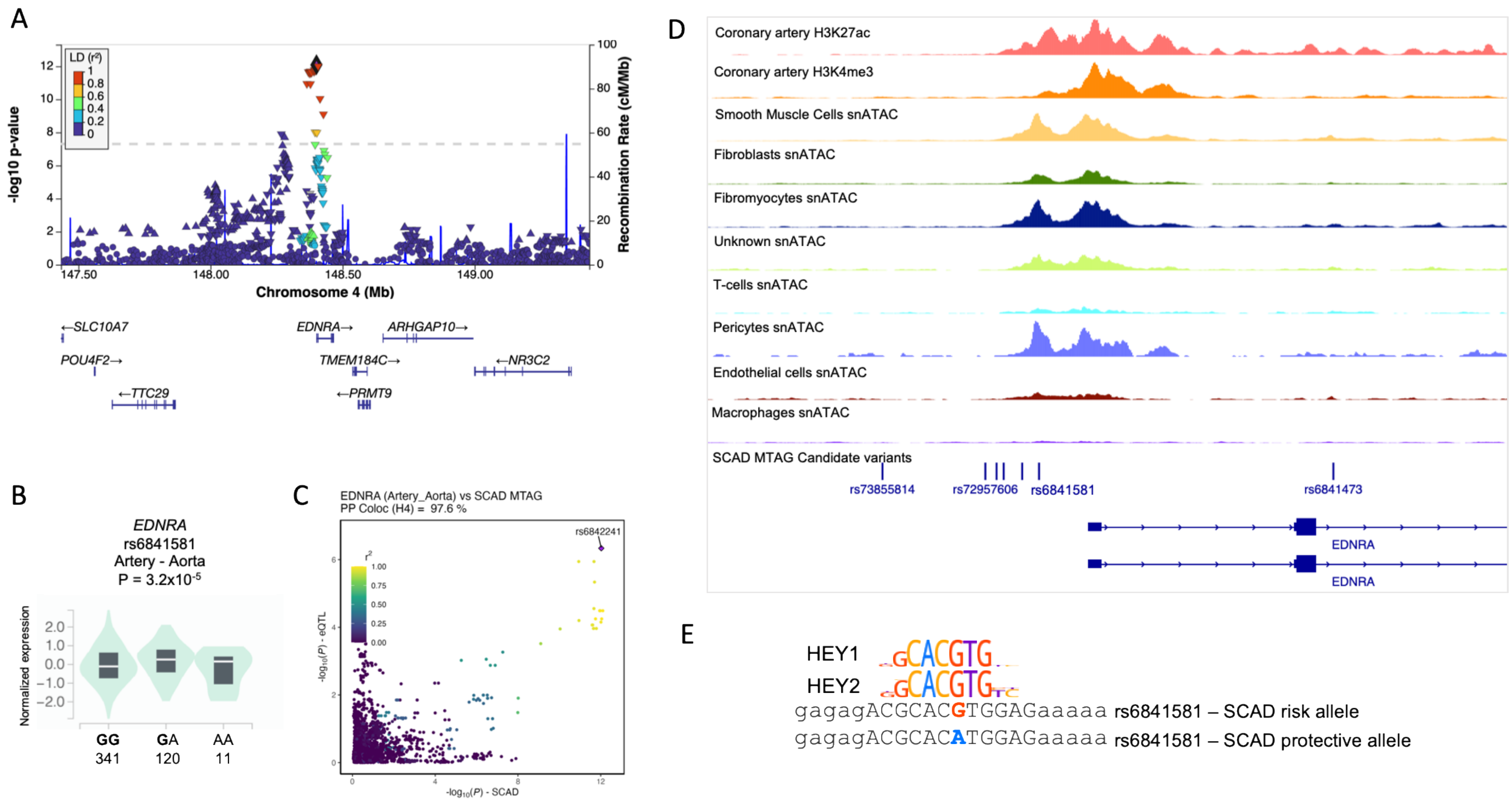


**Supplementary Figure 6. Functional annotation at *EDNRA* locus.**

(A) Regional association plot for the chromosome 4 locus at proximity of *EDNRA* from the SCAD MTAG analysis. The upper panel displays –log10 P values on the left y-axis for variants (triangles) across genomic position on chromosome 4 (x-axis, Mb). The lead variant rs6841581 is indicated by a black-lined purple diamond, and other variants are coloured according to their LD ($r^2$) with the lead SNP, as shown in the legend. The local recombination rate is overlaid as a line referenced to the right y-axis. The lower panel shows RefSeq gene models with gene symbols and transcriptional orientation. (B) Violin plot of *EDNRA* normalized expression by genotype of variant rs6841581 in Aorta from GTEx v10. The number of samples per genotype is shown below the x-axis, with SCAD risk allele in bold. cis-eQTL P-value is indicated over the plot. (C) Scatterplot representation of SCAD MTAG association (*x*-axis) versus *EDNRA* Aorta eQTL association (*y*-axis), showing genetic colocalization of both signals. Posterior probability of two signals to share a causal variant is indicated over the graph (97,6%). Dot colour indicates LD ($r^2$) with rs6842241, identified as the most likely variant to cause association with both traits. (D) Genome-browser view focusing on SCAD candidate variants at proximity of *EDNRA* promoter on chromosome 4. Tracks show Coronary artery H3K27ac and H3K4me3 ChIP-Seq as well as single-nucleus chromatin accessibility (snATAC-seq) in top cellular clusters from coronary artery snATAC analysis. Lead variant rs6841581 directly overlaps a region accessible in SMCs, Fibromyocytes and Pericytes. (E) Representation of genetic sequence at proximity of rs6841581, with SCAD risk allele G corresponding to HEY1/2 canonical binding site (logo representation).

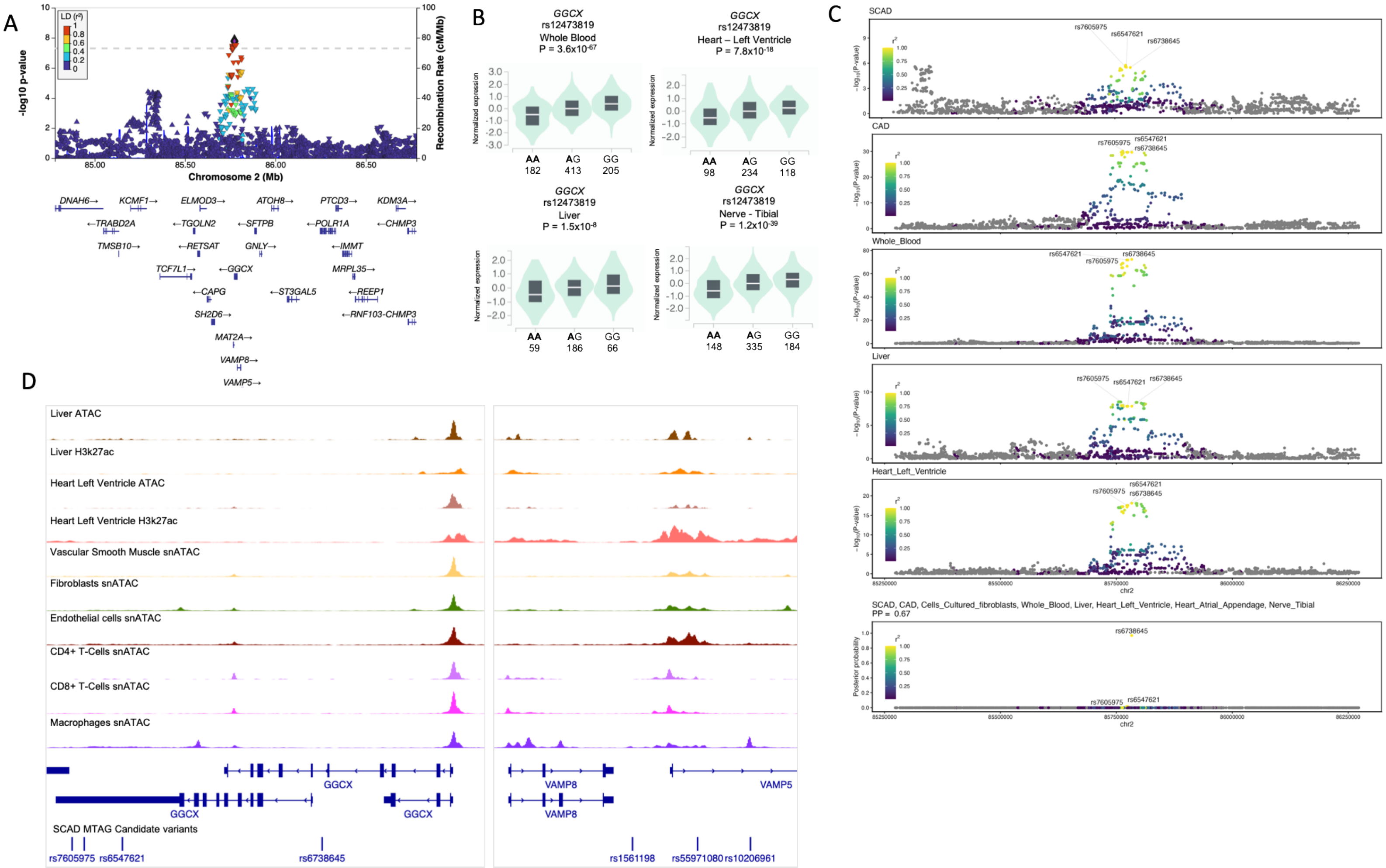


**Supplementary Figure 7. Functional annotation at *GGCX* locus.**

(A) Regional association plot for the chromosome 2 locus at proximity of *GGCX* from the SCAD MTAG analysis. The upper panel displays –log10 P values on the left y-axis for variants (triangles) across genomic position on chromosome 2 (x-axis, Mb). The lead variant rs12473819 is indicated by a black-lined purple diamond, and other variants are coloured according to their LD ($r^2$) with the lead SNP, as shown in the legend. The local recombination rate is overlaid as a line referenced to the right y-axis. The lower panel shows RefSeq gene models with gene symbols and transcriptional orientation. (B) Violin plots of *GGCX* normalized expression by genotype of variant rs12473819 in four tissues from GTEx v10 (Whole Blood, Heart Left Ventricle, Liver, Tibial Nerve). The number of samples per genotype is shown below the x-axis, with SCAD risk allele in bold. cis-eQTL P-value is indicated over the plots. (C) Regional association plots at *GGCX* locus for SCAD GWAS, CAD GWAS and eQTL association with *GGCX* expression in Whole Blood, Liver and Heart Left Ventricle. Multi-trait colocalization was performed and indicated a colocalization of genetic associations with these traits and additional eQTL associations (Heart Atrium, Tibial Nerve) with a high degree of probability (67%). Lower panel shows the colocalization score for each variant. Dot colour indicates LD ($r^2$) with rs6738645, identified as the most likely variant to cause association with all traits. (D) Genome-browser view focusing on SCAD candidate variants at GGCX locus on chromosome 2. Tracks show Liver and Heart Left ventricle H3K27ac ChIP-Seq and chromatin accessibility (ATAC-seq) as well as single-nucleus ATAC signal in six cellular clusters from a multi-tissue snATAC analysis, corresponding to immune and vascular clusters present in a wide variety of tissues. Top variants from multi-trait colocalization analysis overlap GGCX 3'UTR and intronic regions (left panel). Other SCAD candidate variants overlap two open chromatin regions located in *VAMP5* first intron, active in vascular smooth muscle cells, endothelial cells and macrophages.
